# COVID-19 Pandemic Impact on Sexually Transmitted Infection Testing in a College Setting

**DOI:** 10.1101/2021.10.08.21264716

**Authors:** Agustina Marconi, Elizabeth C Falk-Hanson, Megan Crass, Peter Campbell

## Abstract

**Objective:** Assess the impact of the pandemic on STI (sexually transmitted infections) testing in a college health setting.

**Design:** Exploratory analysis of the number of STI tests done, positive rates for those tests and of percentage of “compliance to follow-up” from March to December 2020 and its comparison with historical data at the University Health Services, UW-Madison.

**Sample:** students’ STI tests during the analyzed period.

**Measurement:** Observed (2020) vs Expected (2015-2019, average) number of STI tests, positive rate, compliance to follow-up testing for STIs.

**Results:** There was a significant decrease in the number of tests done and increase of positive rate when compared to historical for total sample and per sex. There was a decrease in the percentage of follow-up for the entire sample and females and an increase for males.

**Conclusions:** Considering the three outcomes assessed, we observe an impact in STI testing during the pandemic. In concordance with national data, our analysis shows significant declines in STI testing and follow-up during 2020 compared to previous years and an increase in positivity rate. The finding of higher positivity with lower number of tests is likely due to triaging patients, facilitating testing for those at highest risk of infection.

## Introduction

### Health Care Systems during Pandemic

The COVID-19 pandemic has had an extraordinary impact on worldwide health and health systems, challenging local and global capacities to respond.^1^ The implications significantly altered aspects of daily life and health care access, including routine health services.^2^

Due to limited intensive care resources, efforts focused primarily on protecting access to critical care services. Preventive services were reduced, and many non-urgent visits were significantly delayed or changed to virtual platforms to preserve personal protective equipment supply and limit patients’ and clinical staff’s exposure risks. Many people avoided seeking routine and even urgent health care, including nearly 60% of young adults aged 18-24 in one large survey. Literature also demonstrates decreased outpatient visits provided in 2020, especially during March-May. ^3, 4^ As it became evident that the pandemic would last longer than a few weeks or months, it was clear that primary health care, including sexual health services, could not be indefinitely delayed without risking harm.

### Sexually transmitted infections (STIs)

All time high cases of chlamydia and gonorrhea were reported in 2019 in the United States, and more than half of these occur among people aged 15-24.^5^ Sexually transmitted infection rates are high in college settings, with chlamydia rates comparable to those seen in family planning and prenatal clinics.^6^ The long-term consequences of chlamydia and gonorrhea include a higher risk of HIV acquisition, pelvic inflammatory disease (PID), infertility, and ectopic pregnancy. Screening for chlamydia has demonstrated reduced rates of PID among women.^7^

While the orders to socially distance implemented to limit COVID-19 spread should have similarly limited spread of STIs, it is unknown to what extent people were following the recommendations as it pertained to sexual activity.^8^ For some young people, these guidelines likely reduced partnered sexual activity.^9,10^ In contrast, other studies found that sexual activity level stayed stable or increased, including with new partners or sex outside of one’s home.^11^

Early during the pandemic, surveillance systems showed a decrease in the rate of STIs when compared to historical data^12^, whereas other settings found that despite lockdown orders, rates were similar.^13^ Given the known incidence and potential sequelae of untreated infections, access to high quality sexual health services is a key focus of our student health center.

### UW-Madison. University Health Services (UHS). STI-related services

The University of Wisconsin-Madison (UW) is a large public research university in the Midwest United States with a total fall 2020 enrollment of 45,540.^14^ In 2020, 69.5% of enrolled students were undergraduates, 52.2% were female, 12.9% were international students, and 65% were white.^15^ University Health Services (UHS) is a college health clinic serving UW-Madison students. There are multiple medical departments which provide STI testing. In 2017, UHS developed a protocol to assess compliance to follow up STI testing, showing significant differences within studied groups.^16^

Early in the pandemic, it became apparent that UHS needed to balance the risks of undetected STIs and the risks to staff, patients, and the community associated with having traditional in-person visits. Self-testing lab only appointments have been available since 2017. These were only for females due to limitations with the electronic health record and clinical concern about needing to support extragenital site testing for males with male sexual partners (MSM), in whom only performing genital site testing may miss half or more of infections.^17,18,19^ A team was working on expanding this to be inclusive but had not yet implemented changes.

Up until March 13, 2020, clinical access at UHS was normal. Afterward, all visits started as phone visits in order to triage. While the CDC offered guidance for STI management with limited clinical interactions in their April 6th, 2020 Dear Colleague Letter^20^, it was quickly noted by clinicians that there was an urgency to establish a safe means of testing our patients. Testing for STIs was deemed appropriate for in-person care by late March, particularly for patients with symptoms or higher risks, but this still required a telephone assessment. A team of UHS staff representing informatics and clinicians coordinated to expand web-booking of STI self-testing to all genders. This was released April 10, 2020 and complemented by phone triage for people experiencing symptoms or anyone recommended to complete extragenital site or additional screening.

Life on campus was dramatically different during this time period. Students were advised to move out of dorms during spring break and all classes moved to virtual, effective March 23, 2020. Classes remained online through summer with a mix of online and in-person classes during the majority of fall semester, with a return to online for the final weeks of fall semester 2020.

According to unpublished internal campus data, it is estimated between 16,457 and 19,751 students remained on campus or the surrounding areas and were potential users of UHS services during this time. Historically, over 45,000 students are enrolled per year at UW-Madison and eligible to utilize UHS. Per state guidelines, as well as for internal quality improvement initiatives, positive tests for chlamydia and gonorrhea are routinely tracked. Given all the changes to lifestyle and clinical access, we wished to compare STI testing for 2020 following declaration of the global pandemic in March to preceding years. The goal of the present study is to assess the impact of the pandemic on STI testing in a college health setting. To do this we assessed:

1. Number of STI tests done during pandemic months in 2020 compared to historical data and
2. Positive rates for STI during pandemic months, and compared them with historical data and
3. Percentage of compliance to STI follow up during pandemic months and compared them with historical data.

## Methods

### Design

We developed an exploratory analysis of the number of STI tests done, positive rate for STI tests and of the percentage of “compliance to follow up to STI” for Primary Care, Sexual Health and Gynecology visits from March to December 2020 and its comparison with historical data (2015-2019) in college students at the University Health Services (UHS), UW-Madison. For the design and statistical comparison, we adapted the World Health Organization (WHO) recommendations for rapid mortality surveillance and epidemic response for a morbidity event like STl.^21^ For the analysis we used total numbers of STI tests, positive rate for STI and percentage of compliance to follow up for the total period and per month and in the same period in the past 5 years (2015-2019). We compared the observed 2020 number of STI tests done, positive rate for STI and percentages of compliance to STI follow up with the expected number of STI tests done, positive rate for STI and percentage of compliance to STI follow up from historical data. We checked the relative change (X-Xhistorical/Xhistorical) as percentage above or below baseline and the higher limit of the 95% confidence interval (CI 95%). We reported total data for number of STI tests done, positive rate for STI and percentage of compliance and we then assessed female and male reports separately for all three outcomes. If a certain month in 2020 had three or less registers, we took a conservative approach and selected the historical worst follow up scenario for the same month. An Institutional Review Board (IRB) approved the protocol.

To measure number of tests done, positive rate, and compliance to follow up testing for STIs during the COVID-19 pandemic, we used:

1. Expected number of STI tests, positive rate, compliance to follow up testing for STIs to have occurred on a monthly basis in the same period in the past 5 years (based on historical data 2015-2019), and
2. Number of STI tests, positive rate, compliance to follow up testing for STIs that have occurred/observed in the analyzed period (2020)

#### Definitions

- Sexually transmitted Infection (STI). For this analysis we will utilize the most commonly diagnosed infections, chlamydia and gonorrhea, as the total STI. We excluded other diagnoses, including HIV, HSV, HPV, and syphilis.
- Positive rate STI: total positive STI tests/total STI tests done in a period of time.
- Compliance to follow up testing for STIs: is defined as repeating a test after 4 to 26 weeks of first treatment. This is based upon CDC guidance to repeat testing 3 months after treatment, with opportunistic screening when that person next presents for care.^7^ The compliance to follow up is related to tests and not to individuals as people can test positive for chlamydia or gonorrhea several times a year. It is measured as a percentage.

## Results

Table 1 shows the number of STI tests done between March and December 2020 and its comparison with historical data. The number of STI tests done in the studied period was 5501. This is a significant decrease of 51.2% when compared to the 11280 average tests from 2015-2019. There is a significant decrease when comparing 2020 number of tests with the historical upper Confidence Interval 95% for the total period (−58.1%). All analyzed months show a significant decrease in the number of tests when compared to historical baseline average with the worst scenario being April (−89%), May (−75.8%) and September (−53.4%). The comparisons to the upper CI 95% were also significant for all 10 months analyzed. When stratifying by sex, both females and males have a similar significant decrease in the total period of -51.9% and -51.6% respectively as well as a significant decrease when comparing with historical upper CI 95% (−58.7% for females and -57.9% for males). All months had a significant decrease in the number of tests both for females and males.

**Table 1.** STI tests done during Covid-19 Pandemic and historical comparison. Total and stratified by per sex and months. UHS. UW- Madison, March-December 2020.

|  | Month | 2015-2019 Average<br>monthly tests (95%<br>CI) | 2020 tests done (n) | % below baseline | % below<br>threshold |
| --- | --- | --- | --- | --- | --- |
| Total | March | 1219 (1060-1378) | 1048 | -14.0 | -23.9 |
|  | April | 1374 (1081-1668) | 151 | -89.0 | -90.9 |
|  | May | 1008 (855-1160) | 244 | -75.8 | -79.0 |
|  | June | 514 (466-562) | 269 | -47.6 | -52.1 |
|  | July | 591 (524-658) | 300 | -49.2 | -54.4 |
|  | August | 775 (653-898) | 434 | -44.0 | -51.7 |
|  | September | 1624 (1313-1935) | 757 | -53.4 | -60.9 |
|  | October | 1617 (1248-1985) | 865 | -46.5 | -56.4 |
|  | November | 1347 (1038-1385) | 857 | -36.4 | -45.3 |
|  | December | 1211 (1038-1385) | 576 | -52.5 | -58.4 |
|  | Total | 11280 (9437-13123) | 5501 | -51.2 | -58.1 |
| Female | March | 752 (674-829) | 638 | -15.1 | -23.1 |
|  | April | 876 (692-1061) | 90 | -89.7 | -91.5 |
|  | May | 578 (469-688) | 145 | -74.9 | -78.9 |
|  | June | 283 (258-308) | 146 | -48.4 | -52.6 |
|  | July | 333 (281-384) | 181 | -45.6 | -52.9 |
|  | August | 459 (379-539) | 244 | -46.9 | -54.8 |
|  | September | 1032 (812-1252) | 446 | -56.8 | -64.4 |
|  | October | 1045 (806-1283) | 606 | -42.0 | -52.8 |
|  | November | 879 (741-1017) | 524 | -40.4 | -48.5 |
|  | December | 743 (652-834) | 338 | -54.5 | -59.5 |
|  | Total | 6981 (5822-8140) | 3358 | -51.9 | -58.7 |
| Male | March | 460 (380-540) | 394 | -14.3 | -27.0 |
|  | April | 486 (380-592) | 59 | -87.9 | -90.0 |
|  | May | 423 (370-477) | 96 | -77.3 | -79.9 |
|  | June | 228 (194-262) | 123 | -46.0 | -53.0 |
|  | July | 257 (231-282) | 116 | -54.8 | -58.9 |
|  | August | 309 (272-346) | 177 | -42.7 | -48.8 |
|  | September | 576 (484-668) | 299 | -48.1 | -55.2 |
|  | October | 558 (434-681) | 241 | -56.8 | -64.6 |
|  | November | 455 (379-531) | 294 | -35.4 | -44.7 |
|  | December | 453 (378-529) | 234 | -48.4 | -55.7 |
|  | Total | 4205 (3581-4828) | 2033 | -51.6 | -57.9 |

Created by the authors

Table 2 shows the positive rate of STI tests done in the analyzed period and its comparison with historical data. For the whole 2020 period, the positive rate was 4.2%, while the historical positive rate was 3.2%. This 30.8% increase was significant when comparing it to baseline and when compared to the historical upper CI 95% (18.5%). When stratified by sex, both females and males have a similar positive rate, 4.2% and 4.5% respectively. Both are significant increases in positive rate from baseline (46.1% and 18.5%). There is also a significant increase when comparing positive rate with historical upper CI 95% for the total period (28.5% and 7.1% respectively). When analyzing data per month, female students have a significant increase in positive rate for STI in April, May, July, September and November and a significant decrease in July, October and December. In male students, April, May, September, November and December were significantly above historical positive rate and June, August and October were significantly below historical positive rate.

**Table 2.** STI tests positive rates during Covid-19 Pandemic and historical comparison. Total and stratified by per sex and months. UHS. UW- Madison, March-December 2020.

|  | Month | 2015-2019 Monthly average (95% CI) | 2020 percentage of positivity x 100 | % above/below baseline | % above/below threshold |
| --- | --- | --- | --- | --- | --- |
| Total | March | 3 (2.4-3.4) | 3.2% | 9.0 | -8.4 |
|  | April | 3 (2.2-3.8) | 6.0% | 96.4 | 55.0 |
|  | May | 2.8 (2.5-3.2) | 4.1% | 44.5 | 28.6 |
|  | June | 3.7 (2.9-4.4) | 4.5% | 22.0 | 1.7 |
|  | July | 3.5 (2.5-4.5) | 3.7% | 3.0 | -19.9 |
|  | August | 3.3 (2.3-4.3) | 3.0% | -9.0 | -30.0 |
|  | September | 3.1 (2.6-3.6) | 4.6% | 48.1 | 27.5 |
|  | October | 3.4 (2.6-4.1) | 2.7% | -22.0 | -36.3 |
|  | November | 3(2.3-3.7) | 5.8% | 94.7 | 57.3 |
|  | December | 3.1 (3-3.3) | 4.3% | 38.8 | 33.4 |
|  | Total | 3.2 (2.9-3.5) | 4.2% | 30.8 | 18.5 |
| Female | March | 2.5 (1.8-3.1) | 2.7% | 8.1 | -14.4 |
|  | April | 2.9 (2.1-3.6) | 6.7% | 132.7 | 85.1 |
|  | May | 2.6 (2.0-3.6) | 4.1% | 61.5 | 31.3 |
|  | June | 2.8 (1.8-3.7) | 5.5% | 97.0 | 46.6 |
|  | July | 3.3 (2.1-4.5) | 3.3% | -0.9 | -27.1 |
|  | August | 3.1 (2.0-3.5) | 4.1% | 33.2 | -2.2 |
|  | September | 3.0 (2.5-3.4) | 4.3% | 43.8 | 25.2 |
|  | October | 2.8 (2.0-3.5) | 2.6% | -4.8 | -25.4 |
|  | November | 2.9 (2.3-3.5) | 6.1% | 111.0 | 74.7 |
|  | December | 3.1 (2.5-3.6) | 2.7% | -13.0 | -25.9 |
|  | Total | 2.9 (2.5-3.3) | 4.2% | 46.1 | 28.5 |
| Male | March | 3.9 (3.3-4.5) | 4.3% | 10.7 | -3.8 |
|  | April | 3.4 (2.2-4.6) | 5.1% | 49.9 | 10.7 |
|  | May | 3.2 (2.6-3.8) | 4.2% | 30.6 | 9.6 |
|  | June | 4.8 (4.4-5.3) | 3.3% | -32.9 | -38.6 |
|  | July | 3.9 (2.3-5.4) | 4.3% | 10.9 | -20.5 |
|  | August | 3.7 (2.8-4.6) | 2.3% | -38.4 | -50.5 |
|  | September | 3.5 (2.6-4.3) | 5.4% | 53.9 | 24.1 |
|  | October | 4.7 (3.7-5.6) | 2.9% | -37.9 | -48.5 |
|  | November | 3.3 (2.1-4.4) | 6.1% | 86.7 | 38.3 |
|  | December | 3.3 (2.7-4.0) | 6.8% | 104.8 | 73.0 |
|  | Total | 3.8 (3.4-4.2) | 4.5% | 18.5 | 7.1 |

Created by the authors

Table 3 and chart 1 show the analyzed data for compliance to follow up for the whole sample and for females and males. Compliance for March-December 2020 was 53%, whereas historical compliance was 58.8%. This means a significant decrease in the percentage of follow up for the whole period of -4.7% with a significant decrease when compared to the historical upper CI 95% (−9.5%). In the analysis per month, we observe a significant decrease of the percentage of follow up in March, April, May, July and November. On the other hand, we observe significant increase in compliance to follow up in October and December. Compliance for March-December 2020 for females was 54%, whereas the historical percentage of compliance for the whole period in females was 62%. This means a significant decrease in the percentage of follow up for the whole period of -11.5% with a significant decrease when compared to the historical upper CI 95% (−16%). In the analysis per month for females’ compliance, we observe a significant decrease of the percentage of follow up in March, April, June, July, November and December. On the other hand, we observe no significant increase or decrease in compliance to follow up for the rest of the months. When analyzing percentage compliance for March-December 2020 for males, we obtain 53.7%, whereas the historical percentage of compliance for the whole period in males was 48.3%. This means a significant increase in the percentage of follow up for the whole period of 11.2% with a significant increase when compared to the historical upper CI 95% (2.2%). In the analysis per month, we observe a significant increase of the percentage of follow up for males in March, April, October and December. On the other hand, males’ compliance to follow up significantly decreased in May, July and August.

**Chart 1:**
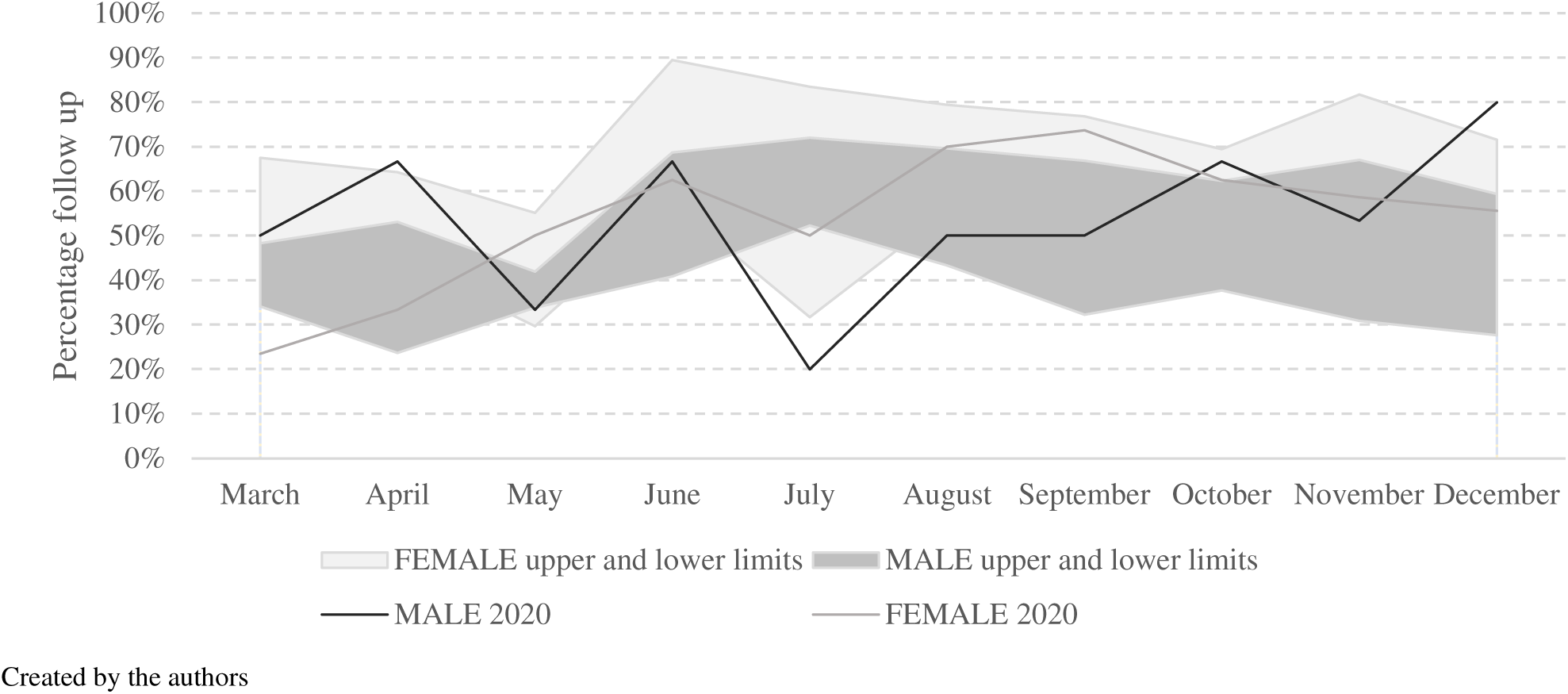
Percentage of STI follow up by sex, March-December 2020 and compared to the upper and lower limits of historical follow up (95% CI). Medical Services UHS, UW-Madison.

**Table 3.**
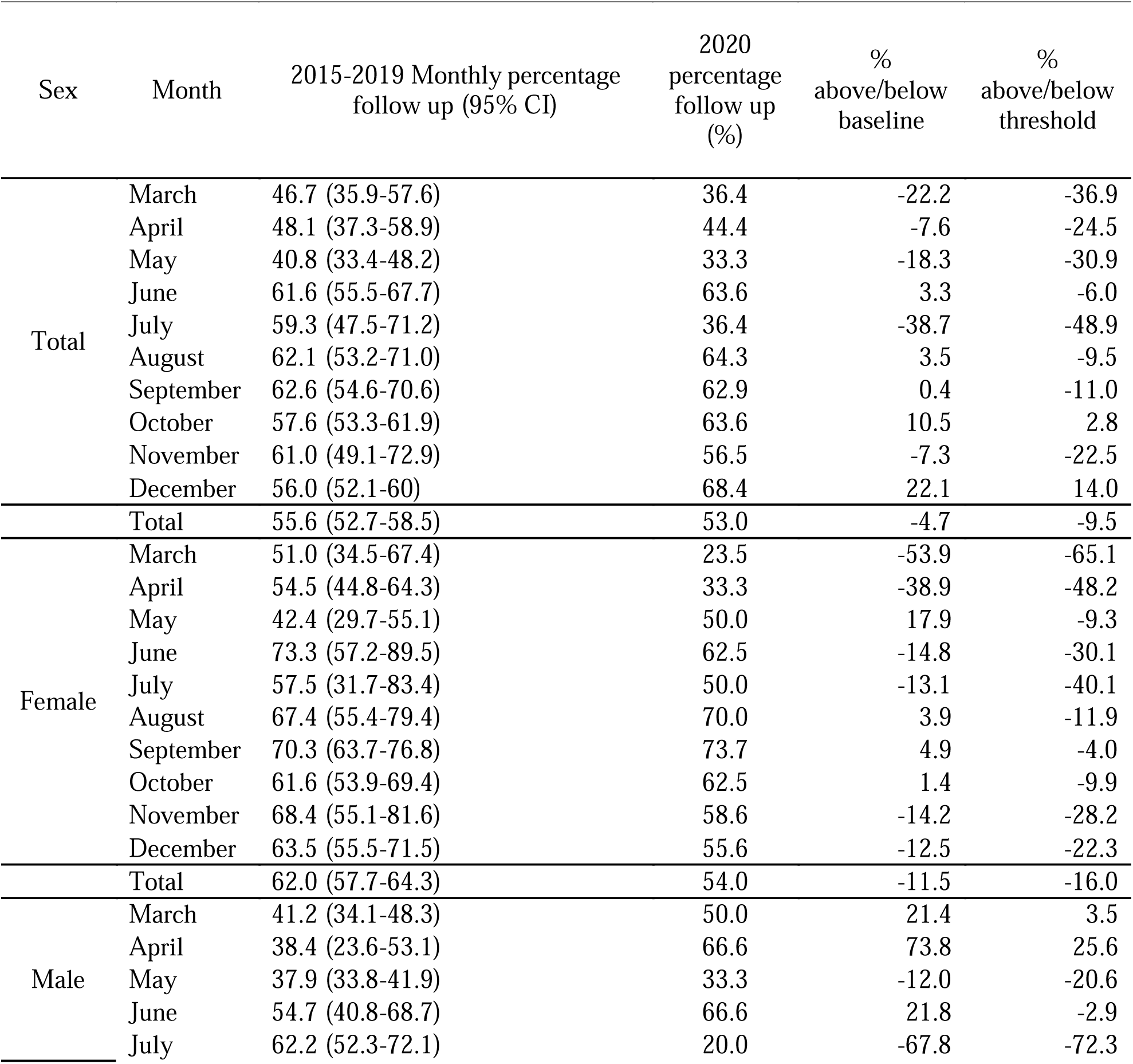
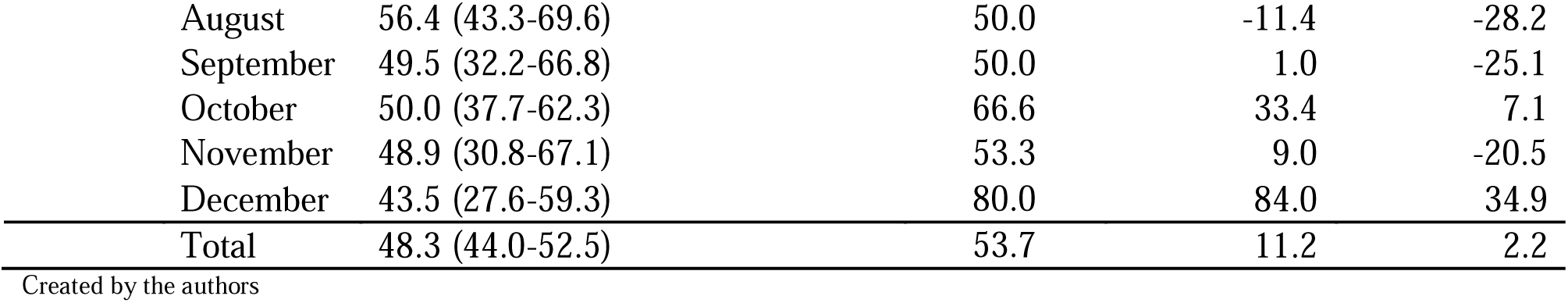
STI tests percentage of follow up during Covid-19 Pandemic and historical comparison. Total and stratified by per sex and months. UHS. UW- Madison, March-December 2020.

| Sex | Month | 2015-2019 Monthly percentage follow up (95% CI) | 2020 percentage follow up (%) | % above/below baseline | % above/below threshold |
| --- | --- | --- | --- | --- | --- |
| Total | March | 46.7 (35.9-57.6) | 36.4 | -22.2 | -36.9 |
|  | April | 48.1 (37.3-58.9) | 44.4 | -7.6 | -24.5 |
|  | May | 40.8 (33.4-48.2) | 33.3 | -18.3 | -30.9 |
|  | June | 61.6 (55.5-67.7) | 63.6 | 3.3 | -6.0 |
|  | July | 59.3 (47.5-71.2) | 36.4 | -38.7 | -48.9 |
|  | August | 62.1 (53.2-71.0) | 64.3 | 3.5 | -9.5 |
|  | September | 62.6 (54.6-70.6) | 62.9 | 0.4 | -11.0 |
|  | October | 57.6 (53.3-61.9) | 63.6 | 10.5 | 2.8 |
|  | November | 61.0 (49.1-72.9) | 56.5 | -7.3 | -22.5 |
|  | December | 56.0 (52.1-60) | 68.4 | 22.1 | 14.0 |
|  | Total | 55.6 (52.7-58.5) | 53.0 | -4.7 | -9.5 |
| Female | March | 51.0 (34.5-67.4) | 23.5 | -53.9 | -65.1 |
|  | April | 54.5 (44.8-64.3) | 33.3 | -38.9 | -48.2 |
|  | May | 42.4 (29.7-55.1) | 50.0 | 17.9 | -9.3 |
|  | June | 73.3 (57.2-89.5) | 62.5 | -14.8 | -30.1 |
|  | July | 57.5 (31.7-83.4) | 50.0 | -13.1 | -40.1 |
|  | August | 67.4 (55.4-79.4) | 70.0 | 3.9 | -11.9 |
|  | September | 70.3 (63.7-76.8) | 73.7 | 4.9 | -4.0 |
|  | October | 61.6 (53.9-69.4) | 62.5 | 1.4 | -9.9 |
|  | November | 68.4 (55.1-81.6) | 58.6 | -14.2 | -28.2 |
|  | December | 63.5 (55.5-71.5) | 55.6 | -12.5 | -22.3 |
|  | Total | 62.0 (57.7-64.3) | 54.0 | -11.5 | -16.0 |
| Male | March | 41.2 (34.1-48.3) | 50.0 | 21.4 | 3.5 |
|  | April | 38.4 (23.6-53.1) | 66.6 | 73.8 | 25.6 |
|  | May | 37.9 (33.8-41.9) | 33.3 | -12.0 | -20.6 |
|  | June | 54.7 (40.8-68.7) | 66.6 | 21.8 | -2.9 |
|  | July | 62.2 (52.3-72.1) | 20.0 | -67.8 | -72.3 |

|  |  |  |  |  |
| --- | --- | --- | --- | --- |
| August | 56.4 (43.3-69.6) | 50.0 | -11.4 | -28.2 |
| September | 49.5 (32.2-66.8) | 50.0 | 1.0 | -25.1 |
| October | 50.0 (37.7-62.3) | 66.6 | 33.4 | 7.1 |
| November | 48.9 (30.8-67.1) | 53.3 | 9.0 | -20.5 |
| December | 43.5 (27.6-59.3) | 80.0 | 84.0 | 34.9 |
| Total | 48.3 (44.0-52.5) | 53.7 | 11.2 | 2.2 |

Created by the authors

## Discussion

Considering the three outcomes assessed, we do observe an impact in STI testing during the pandemic. The impact of the safety measures implemented to adapt to the new reality show consequences in the number of visits. In concordance with national data, our analysis shows significant declines in chlamydia and gonorrhea testing during 2020 compared to values from the previous five years^22^. The total number of chlamydia and gonorrhea tests from March to December 2020 significantly decreased when compared to historical data for the total period and in each analyzed month. (Table and Figure 1). Unpublished UHS data shows that from March to December 2020, the total number of visits for Primary Care, Sexual Health and Gynecology decreased 39% during 2020 compared to historical data. Clinical access tightened up to limit in-person visits during times of increased COVID-19 activity on campus, which may have impacted follow-up rates. While it is beyond the scope of this study, a comparison of follow up rates vs COVID-19 activity level on campus may further our understanding of monthly differences.^23^

Overall, given the changes in access to routine clinical health services based on clinic limitations as well as studies demonstrating hesitancy on behalf of patients related to COVID-19, the finding of reduced testing is expected.

Positive STI rates increased for both males and females.^22^ When analyzed by sex, historical data shows the percentage of positivity was higher in male students, consistent with males more often testing because of symptoms.^24^ Additionally, the finding of higher positivity with lower number of tests overall is likely due to triaging of patients and recommendation and facilitation of testing for those at highest risk of infection, including those with symptoms, known contact, or sexual behaviors associated with higher risk for infections. While appearing that we had a more efficient means of testing given the increased positivity rate in setting of reduced testing overall, it is likely that by adhering to the triaging, which was in line with CDC guidance at the time, many asymptomatic infections were missed. Many of these screens often occur during the context of wellness visits, which were primarily completed through telehealth at UHS during this time period. These asymptomatic infections are especially common in women, as well as typical of extragenital site infections, which make up the majority of chlamydia and gonorrhea infections in MSM.^7^ These infections can be associated with serious outcomes, such as PID and infertility in women and an increased risk of HIV acquisition, in particular for rectal infections, in MSM.^7^ While CDC guidance supported empiric treatment of symptomatic illnesses suggestive of STIs, there was little guidance for identifying the common asymptomatic infections that make up the bulk of infections.^25,7^ UHS prioritized ongoing care for patients utilizing HIV pre-exposure prophylaxis (PrEP) services throughout the pandemic, including access to chlamydia and gonorrhea screening. The use of PrEP is advised for people at higher risk for HIV acquisition, and thus more likely to acquire chlamydia and gonorrhea as well, so this may have additionally impacted the positivity rate. The majority of people who use PrEP at UHS are males, which could have particularly impacted the male positivity rate.^7^ Finally, there was a minimal decrease in follow up for the whole sample in the 2020 months analyzed when compared to historical data. Historical UHS analysis shows male students tend to be less compliant to STI follow up than female students (39.6% vs 60.4%).^16^ Although our analysis shows a slightly lower compliance to follow up for male students in 2020 when compared to females (53.7 % vs 54%), male students show an increase in compliance to follow up when compared to male historical data. While a past UHS analysis demonstrated that males with partners of a different sex have lower STI follow up rates than females, this wasn’t true of males with partners of the same sex or males with partners of same and different sex partners^16^, so the triaging and maintenance of PrEP care during the pandemic may have contributed to a higher rate of follow up in males observed in 2020. This analysis did not include sex of partners for comparison. Our prior analysis did not include symptomatic compared to asymptomatic at time of diagnosis; by favoring detection of symptomatic cases during 2020, it’s possible that this experience may impact the likelihood of STI follow up, though to what degree clinical factors impact repeat testing is unclear from the literature.

### Future implications

Despite many perceived barriers to developing new access opportunities, when the urgency was apparent, UHS brought together a team to expedite a process change to meet the needs of patients and clinicians that was also mindful of community safety and resources. The process allowed us to diagnose and treat many STIs in a significantly altered environment. New follow-up analyses will be needed to check for negative health consequences of missed screening opportunities in the college population. However, multiple options for accessing care may allow people to feel more able to access needed services in a confidential way. Living situations changed greatly during the pandemic and more young adults were once again living with family members^26^, so we have maintained multiple access options to meet various needs.

### Limitations

This is an ecological study; therefore, no directionality or association can be established. Our sample includes only college students, so it may not be generalizable to non-college populations. The study only includes tests done at UHS, so follow up testing done elsewhere is unknown. We were limited to binary sex analysis by sourcing through registrar data. Although we have an estimation of students that could potentially have used UHS during the pandemic, we don’t know exactly how many remained near campus monthly.

## Data Availability

All data produced in the present study are available upon reasonable request to the authors

## Contributorship Statement

- Agustina M. Marconi: Research planning and execution. Development or design of methodology. Adapting of models. Conducting a research and investigation process. writing the proposal. Getting IRB approved. Main role leadership responsibility. Application of statistical analysis.
- Elizabeth C Falk-Hanson: Research planning and execution/First ideas of the proposal. Data collection. Data cleaning (Data curation). Writing the initial draft. Preparation and review of the manuscript. Data presentation. Review different versions
- Megan Crass: Research planning and execution. First ideas of the proposal. Data collection. Data cleaning (Data curation). Writing the initial draft.
- Peter Campbell: Research planning and execution. Produce data reports. Preparation of the manuscript. of statistical mathematical techniques.

## Key messages box

- Our analysis shows significant decline*s* in chlamydia and gonorrhea testing during 2020 compared to historical values.
- Positive STI rates increased for both males and females.
- There was a minimal decrease in follow up for the whole sample in the 2020 months analyzed when compared to historical data.
- Male students show an increase in compliance to follow up when compared to male historical data.

## Acknowledgments

Carol Griggs^v^

Courtney Blomme^vi^

Tamra Dagnon^vii^

## Footnotes

v PHD. Director of Business Services and Operations. UHS. UW-Madison

vi MS, RD. Associate Director of Quality, Evaluation and Assessment at University Health Services at UW-Madison

vii B.S. in Human Relations Prosci® Certified Change Practitioner.

